# Data-Driven Real-Time Strategic Placement of Mobile Vaccine Distribution Sites

**DOI:** 10.1101/2021.12.15.21267736

**Authors:** Zakaria Mehrab, Mandy L. Wilson, Serina Chang, Galen Harrison, Bryan Lewis, Alex Telionis, Justin Crow, Dennis Kim, Scott Spillmann, Kate Peters, Jure Leskovec, Madhav V. Marathe

## Abstract

The deployment of vaccines across the US provides significant defense against serious illness and death from COVID-19. Over 70% of vaccine-eligible Americans are at least partially vaccinated, but there are pockets of the population that are under-vaccinated, such as in rural areas and some demographic groups (e.g. age, race, ethnicity). These unvaccinated pockets are extremely susceptible to the Delta variant, exacerbating the healthcare crisis and increasing the risk of new variants. In this paper, we describe a data-driven model that provides real-time support to Virginia public health officials by recommending mobile vaccination site placement in order to target under-vaccinated populations. Our strategy uses fine-grained mobility data, along with US Census and vaccination uptake data, to identify locations that are most likely to be visited by unvaccinated individuals. We further extend our model to choose locations that maximize vaccine uptake among hesitant groups. We show that the top recommended sites vary substantially across some demographics, demonstrating the value of developing customized recommendation models that integrate fine-grained, heterogeneous data sources. In addition, we used a statistically equivalent Synthetic Population to study the effect of combined demographics (eg, people of a particular race and age), which is not possible using US Census data alone. We validate our recommendations by analyzing the success rates of deployed vaccine sites, and show that sites placed closer to our recommended areas administered higher numbers of doses. Our model is the first of its kind to consider evolving mobility patterns in real-time for suggesting placement strategies customized for different targeted demographic groups. Our results will be presented at IAAI-22, but given the critical nature of the pandemic, we offer this extended version of that paper for more timely consideration of our approach and to cover additional findings.

## 1 Introduction

As of August 2, 2021, at least 70% of American adults aged 18 and older had received at least one dose of a COVID- 19 vaccine Reuters (2021). However, in many subpopulations, including young people, Black people, people of Latinx ethnicity, and in rural areas, the vaccination rate runs far below that UCSF (2021); PBS (2021). Strategies have been devised to address vaccine accessibility – free child care, paid time off for employees, and other financial incentives – but these measures have not proven effective for these under-vaccinated demographic groups.

In order to increase the rate of vaccination among the under-vaccinated or less vaccine-enthusiastic populations, the Virginia Department of Health (VDH) has begun deployment of mobile vaccine distribution sites. These mobile units distribute the one-dose Johnson & Johnson vaccine in order to simplify scheduling and encourage “impulse” vaccinations (i.e., seizing the opportunity to get vaccinated when presented). When VDH started this program, deployment was driven primarily by intuition or educated guesses by local public health officials. However, three important factors were not well-addressed in this deployment strategy: (*i*) each demographic group has its own mobility patterns and may frequent different locations; (*ii*) mobility patterns have been evolving as the lockdown has eased; and (*iii*) the locations for these sites have not been well-publicized. Therefore, a more methodical, real-time deployment plan was needed to maximize uptake among targeted demographic groups, and to increase responsiveness to dynamic mobility patterns.

The success of mobile site placements depends on (*i*) the **accessibility** of these sites for the target populations, (*ii*) their willingness to get vaccinated (**acceptance/hesitancy**), and (*iii*) strategic outreach, or **advertisement**, of these sites. In this paper we focus primarily on **accessibility** and secondarily on **acceptance/hesitancy**. To address the **accessibility** aspect, we propose that areas with high foot traffic from the target demographic groups would be productive locations to place mobile vaccination units. To this end, we employ a *dynamic, data-driven* recommendation model, using heterogeneous data sources, that can recommend locations with high probability of vaccination uptake success in *real-time*.

Our model works as follows:

*First*, it uses aggregated and anonymized mobility data from SafeGraph to identify areas with high mobility concentrations and the Census Block Groups (CBGs) that contribute to that traffic. Such data has been used extensively as a means to study the spread of COVID-19 and to track the degree of compliance with social distancing directives Badr et al. (2020); Buckee et al. (2020); Warren and Skillman (2020); Wellenius et al. (2020); Chang et al. (2021a); Wang et al. (2020). The candidate areas are defined as tessellations indexed by Google’s S2 Geometry.

*Second*, it leverages multiple data sources containing the demographic profile of each CBG to adjust the previously computed mobility for target demographic groups using a set of equations. Based on this adjusted mobility, it ranks the tessellations separately for each demographic group.

*Third*, it is equipped with a module to estimate vaccine **acceptance** across different demographic groups in order to refine the previous rankings.

Overall, our model can be described as a rule-based system which consists of a set of rules (equations) used to process the heterogeneous knowledge graph data sources (SafeGraph + Census data) and takes subsequent actions (recommendations). The model also champions fairness and equity, which are lingering issues in AI. Machine learning models can suffer from bias, serving certain demographic groups better than others Sweeney et al. (2019), but our model mitigates this issue by employing the rules after taking into account the racial heterogeneity of CBGs.

To evaluate how our placement strategy compared with actual placements made by public health officials, we review existing mobile distribution site placements from data provided by VDH and observe how many people were vaccinated at these locations. Furthermore, we analyze the robustness of our model for various demographic groups and different weeks.

In line with the continuing support our group has provided to various local, state, and federal public health authorities since the onset of the pandemic, we presented two prototypes of our model to VDH and received valuable guidance integral to the current implementation and the selection of demographic groups for deployment. Our model has been operational since the beginning of June 2021, continuously providing real-time placement recommendations. Although we have not received quantifiable data reflecting the effectiveness of these sites, VDH does rely on these recommendations for mobile vaccination site planning. Francisco Diaz, the Vaccine Administration Support Supervisor for VDH, has stated that this program allows VDH to identify where vaccines are needed. This improves their ability to focus their efforts on reaching target demographic groups in locations that are accessible and convenient to them. His complete statement is available at Diaz (2021).

One of the key aspects where our model outshines complex deep learning models and other location-theory algorithms is its simplicity. For a safety-critical use case such as ours, policymakers prefer a solution that is transparent and easy to interpret. Alternatively, location-theory algorithms are often difficult to solve, while deep learning models are difficult to interpret. Our model alleviates both caveats by incorporating simple rules to generate recommendations and presenting them in a transparent fashion. From this aspect, our model advocates interpretable AI.

In recent times, there have been studies evaluating the effectiveness of different vaccination strategies for different scenarios Saldaña et al. (2021); Chen et al. (2021); Buckner et al. (2021); Jentsch et al. (2021). These studies are mostly conducted by simulating different scenarios using compartmental models, and although they can tell us *who* to vaccinate, the answer to *where* to vaccinate these individuals is still absent in the literature. *Our work is the first of its kind to develop a data-driven model that considers evolving mobility patterns and finds a real-time placement strategy that is accessible to different targeted demographic groups*.

The rest of this paper is organized as follows. We describe similar previous works relying on mobility data in Section 2. In Section 3, we describe the incremental design and implementation of our model to create a ranking of areas for placement of vaccine distribution sites. The delivery process of these recommended areas to VDH is described in Section 4, along with other salient insights about the mobility of different demographic groups obtained from our models and comparisons between our recommended locations and existing sites. Lastly, we explore the utility and implications of our model in Section 5.

## 2 Related Work

### Mobility and Vaccination

A first set of works Saldaña et al. (2021); Chen et al. (2021); Buckner et al. (2021); Jentsch et al. (2021) simulated different vaccination strategies for various scenarios and studied their effectiveness. Jentsch et al. (2021) found that if there is a delay in vaccine availability, it is more effective to target individuals with high social contact instead of focusing on the elderly; Chen et al. (2021) reached similar conclusions, except they compared between high contact people and essential workers. Saldaña et al. (2021) evaluated the effectiveness of different vaccination strategies by simulating a meta-population model across several scenarios. Buckner et al. (2021) used a mathematical model that indicated that the prioritization strategy should vary depending on the objective; for example, targeting essential workers minimizes infection, but targeting older individuals minimizes the number of deaths. In terms of vaccination, these works focus mainly on *who* but not *where*, whereas in our work we focus on both aspects.

In a separate study using anonymized geospatial mobility data, Huang et al. (2021) developed a method for calculating the social contact rate of individuals and assessed the future impacts with and without vaccination. They found that, in the absence of vaccination, it is necessary to strongly enforce physical distancing, and the degree of enforcement should increase in accordance with the population density in the area. They also assessed the impact of vaccination, and inferred that in low-density areas vaccination alone can reduce transmission by a fair amount, but in high-density areas moderate levels of physical distancing are required along with vaccination to achieve the same result.

### COVID-19 systems to aid policymakers

Similar to how our system is designed to provide data-driven, real-time support to policymakers, quite a few systems have been developed to aid policymakers during the pandemic. Some were designed for surveillance purposes, e.g. visualizing infection rates and trends at different spatial resolutions Dong et al. (2020); Peddireddy et al. (2020); Wissel et al. (2020), identifying anomalous hotspots Hohl et al. (2020), and informing policymakers about the necessary levels of restriction in a timely fashion Qiu (2021). Another set of systems was developed to help policymakers observe the effects of different non-pharmaceutical interventions in order to help them make informed decisions Barrett et al. (2007); Beckman et al. (2014); Chang et al. (2021b).

What sets our work apart is that we are the first to develop an operational system that provides weekly updates to policymakers regarding placement strategies for mobile vaccine distribution sites across different demographic groups. Also, unlike other systems which largely focus on surveillance and retrospective analyses, our system provides realtime support to policymakers for mitigating disease transmission.

## 3 Methodology

### 3.1 Datasets

#### Fine-grained mobility data (SafeGraph)

Mobility data can reveal important information about populations, such as where people are visiting and how this behavior evolves over time. For our model, we use anonymized and aggregated data from SafeGraph SafeGraph (2018). It provides detailed information about non-residential locations visited by individuals (e.g. grocery stores, parks), also referred to as *points of interest* (POIs). SafeGraph’s Weekly Patterns dataset^1^, released on Wednesdays with the data for the previous week (Monday through Sunday), includes weekly estimates of visits from CBGs to these POIs. The dataset can be naturally viewed as a bipartite graph, as described later in 3.3. For our work here, we focus on POIs and CBGs in the Commonwealth of Virginia, where visits from 5,293 CBGs to 74,535 POIs were compiled in the latest release.

However, SafeGraph has some limitations. For instance, it does not cover all POIs or populations (e.g., children). Furthermore, depending on the number of devices carried by a user, visits may be underreported or overreported. The GPS signal itself can also be noisy. We describe how we account for some of these limitations in Sections 3.2 and 3.3.

#### Census Data

Since the SafeGraph dataset does not contain demographic information, we use demographic data from the US Census American Community Survey (ACS) in conjunction with the visits from the SafeGraph’s CBG-to-POI data to estimate visits to each POI from each demographic group, or which POIs are frequented by each group. The 2015-2019 release of the 5-year Census data provides information about the populations of different individual demographic groups (e.g. Black, Latinx) at the Census Block Group (CBG) level. The data is presented across different files, each containing information about a set of similar demographic groups. For example, the file *cbg b01* contains populations of different genders across age groups (e.g. Males of 40 To 44 Years, Females of 40 to 44 years) while the file *cbg b02* contains the populations of different races (e.g. Black or African American, Asian). The census data comes with a metadata file called *cbg field description* which contains information about each column across all files and a description about the information each column provides.

One weakness of the Census data is that it does not allow us to produce populations by combining demographic groups from different sets, or groups which are present across different files. For example, there is no obvious way to calculate the population of black males between 40 and 44 years of age from this data. We describe our approach to obtain information about combined demographic groups in the next section.

#### Synthetic Populations Data

As discussed earlier, the Census data does not contain information about the populations of combined demographic groups. To address this problem, we use an approach developed by the Biocomplexity Institute Mortveit et al. (2020) to create a synthetic population of a given region using US Census data and other sources as a base. The synthetic population includes detailed information about each synthetic individual, such as age, race, ethnicity, and income. Given that it is a good representation of the demographic distribution of the actual population, it can be used to address questions about combined demographic groups.

Our basic approach is to take marginals of demographic group *G*_1_ and demographic group *G*_2_ along with Public Use Microdata Sample (PUMS) data to create a joint distribution using *iterative proportional fitting*. Individuals can then be sampled using this joint distribution. A key property of this process is that the marginal distribution of the synthetic data, as well as the joint distribution, is statistically identical to the given input distributions.

#### Vaccination Data

Our final dataset, obtained from VDH, contains the number of individuals per census tract who have received vaccine doses; this data was approved for this study by an Internal Review Board as described in Section 7. However, Safe-Graph and ACS Census data are provided at the CBG level. In order to maintain the same level of resolution across all data sources, we estimate the number of unvaccinated individuals at the CBG level, assuming that the number of doses in a census tract is distributed proportionately across its underlying CBGs.

### 3.2 S2 Geometry

Although SafeGraph provides the number of visits to specific POIs, the resolution level is quite dense for calculating the placement of mobile vaccination sites. Furthermore, the signals picked up for a specific POI do not necessarily indicate that the device was present in that specific POI at that time of collection due to the noise associated with Global Positioning Systems (GPS) signals. In order to address these issues, instead of considering individual POIs with high foot traffic for potential site placement, we identify geographical areas with high foot traffic as described below:

• We divide each county in Virginia into much smaller *areas*, each of which contains a group of POIs.

• For each area, we aggregate the foot traffic for all of the POIs in that area, then rank each area inside a region based on this aggregated foot traffic.

To partition Virginia into regions, we use Google’s S2 geometry^2^, which divides the world map into nested cells of decreasing size (L0 - L30). Level L0 is one cell representing the entire map; it contains 4 L1 cells, each of which contains 4 L2 cells, each of which contains 4 L3 cells, and so on.

### 3.3 Placement model

Our data-driven model was guided by two preceding proof-of-concept prototypes. First, a pilot study was conducted in the Southside Health District of Virginia that recommended mobile vaccination site placement using traditional location allocation methodologies that minimized driving time, but limited the locations to public sites (e.g. fire houses, box stores, and schools). We concluded from this study that areas with high foot traffic may be good candidates for vaccine distribution sites, and, since commercial venues may see higher foot traffic than public sites, it is worth exploring a more diverse set of POIs.

Building on this information, we developed a second prototype model that recommended a set of L14 cells for each L8 cell based on the aggregated foot traffic of the POIs within each L14 cell. These results were presented to VDH, but reconciling the L8s to actual locations required too much overhead for public health officials; thus, VDH requested that we provide recommendations at the locality (city/county) level. Furthermore, areas with the highest foot traffic were not necessarily the ones most visited by the under-vaccinated demographic groups, so VDH requested that we explore recommending areas frequented by specific demographic groups. These recommendations helped us shape our final model, described below. More detailed information about our previous models can be found in the Appendix of this paper.

#### Mobility Network

One input to our placement model is a dynamic mobility network which can be represented as a bipartite graph *G*(*V, E*), where *V* is the set of nodes and *E* is the set of time-varying edges. *V* is the union of two disjoint sets *C* = {*c*_1_, …, *c*_*m*_} and *P* = {*P*_1_, …, *P*_*n*_}. Here *C* represents the set of CBGs, and *P* is the set of POIs in the dataset. Each edge (*c*_*i*_, *p*_*j*_) is associated with a weight 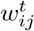, the number of people from *c*_*i*_ visiting *p*_*j*_ at time *t*.

Let *D* = {*d*_1_, *d*_2_, …} be a set of demographic groups of interest. Each CBG *c*_*i*_ is associated with its overall population *N*_*i*_ and the population 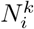, which is specific to demographic group *d*_*k*_ (where *d*_*k*_ ∈ *D*). Each CBG *c*_*i*_ is also associated with *M*_*i*_, the number of mobile devices from *c*_*i*_ captured by SafeGraph. Subsequently, each POI *p*_*j*_ is also associated with a small geographic area denoted by *S*_*j*_ and a large region indicated by *L*_*j*_. In our final model described below, each small geographic area is an L14 cell of Google’s S2 geometry as described in Section 3.2 and each large region is a county. Therefore, we use these notations for denoting the L14 cell and county also in subsequent paragraphs.

Our placement model takes the dynamic bipartite graph generated from SafeGraph and a set of demographic groups of interest, and generates as output a ranked list of areas as potential candidates for setting up mobile vaccination units within each larger region. This is performed as follows:

• First, we adjust the weights along the edges of the graph to estimate the number of actual visits to each POI to mitigate the under-reporting issue discussed in 3.1. Due to this issue, the actual number of visits is higher than the reported number of visits. This issue is addressed by *upweighting* the visits along each edge as : 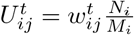

• Second, we adjust the weights for each demographic group *d*_*k*_ by obtaining an apportioned estimate of how many people of that group from CBG *c*_*i*_ visited a POI *p*_*j*_. In other words, we *apportion* the visit along the edge for a particular demographic *d* as : 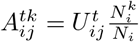

• Third, each area (L14 cell) within a region (county) is ranked based on the apportioned visits to the POIs within that cell. Let 𝒮 = {*s*_1_, *s*_2_, …} be the set of all distinct areas. Then, we *aggregate* the apportioned number of visits to the POIs of a particular L14 cell *s*_*L*_ and calculate its *visit count* at time *t* for a particular demographic group *d*_*k*_ as: 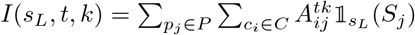

where:

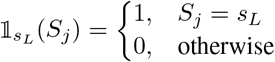

• Finally, we *group* each area (L14 cell) *S*_*j*_ by their corresponding region (county) *L*_*j*_, then sort by their *visit counts*. For each county, the 25 top-ranked areas are reported as candidate sites. The conceptual pipeline for the model is presented in Figure 1a.

**Figure 1:**
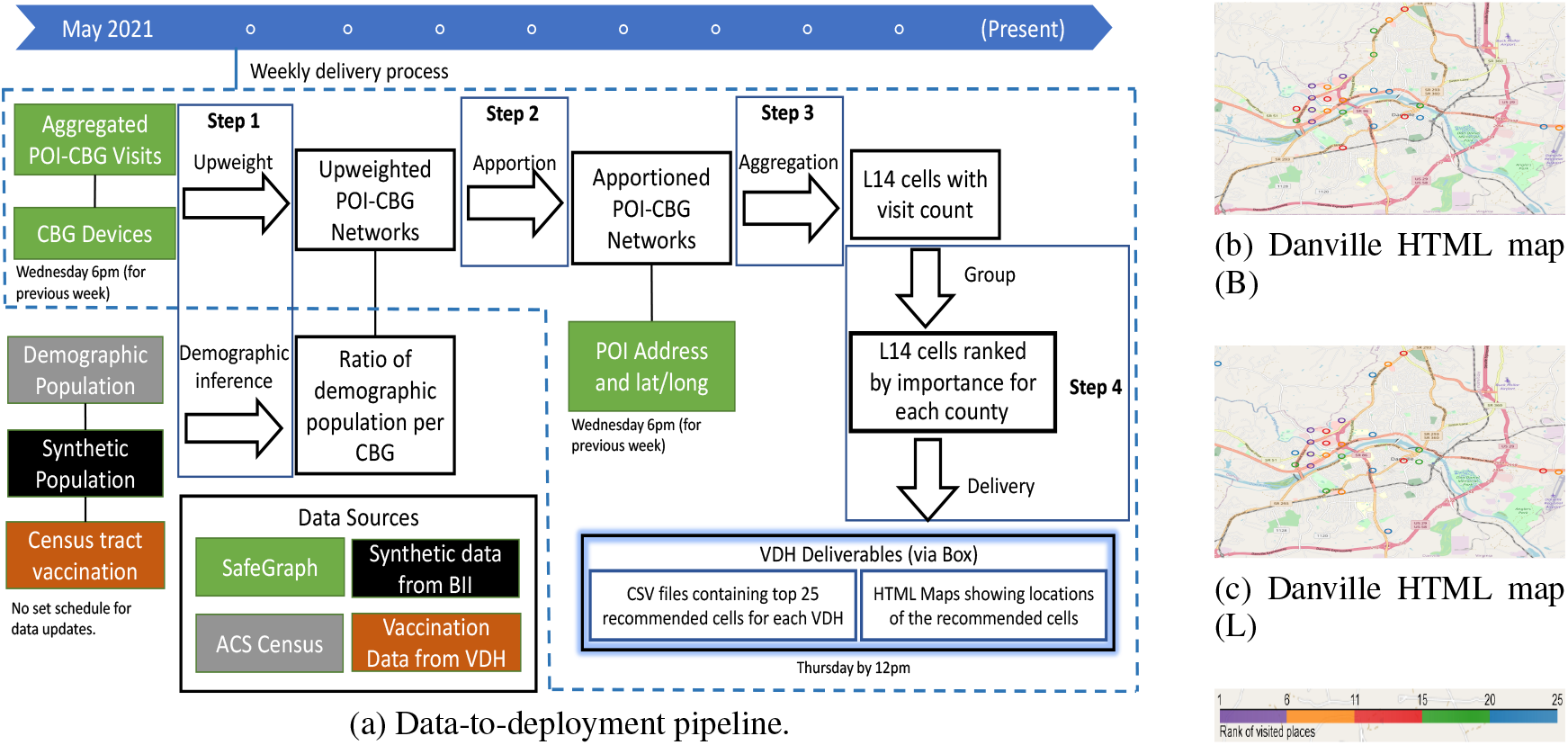
Detailed breakdown of the pipeline of our recommendation model (1a). The pipeline is run weekly on Wednesdays once Safegraph data is updated with the mobility information about the previous week. Sample HTML deliverables generated by the model are shown for Danville City for the week of June 21-June 27 for two demographic groups (1b,1c). The deliverables to VDH are generated by noon on Thursday.

### 3.4 Demographic Acceptance/Hesitancy

Even if vaccination units are placed in easily accessible locations, some individuals may be unwilling or afraid to get vaccinated CNN (2021); PBS (2021). We employ a simulation-based approach to infer the hesitancy level of different demographic groups at the county level; this approach is described in the Appendix of this paper. After the simulation, for each timestamp *t*, each CBG *C*_*i*_ and a demographic group *d*, we have the population count, number of individuals who had at least one dose of vaccine 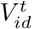, and the estimated number of hesitant individuals as per the calibrated model 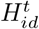. In this section, we describe how we incorporate these values into our model.

Overcoming the hesitancy threshold in areas where vaccine acceptance is low can be a challenge; for this reason, placing mobile sites in these areas may not be the best strategy for maximizing uptake. However, hesitant people may be influenced to take the vaccine if they see their peers getting vaccinated (i.e. peer pressure). Therefore, our approach is to improve accessibility to the vaccine-accepting in areas that may also be frequented by the hesitant. Our method to update the apportioned value in Step 2 of Section 3.3 using this new dataset is described as follows:

• We still pick locations with high foot traffic (emphasis on the original value of the apportioned visits 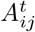)

• We also give higher priority to areas with greater numbers of unvaccinated individuals.

• If vaccine-hesitant people can be influenced by peer pressure, then we want to lower priority in areas where most of the unvaccinated are hesitant.

Based on this, we calculate an updated apportioned weight 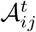 by incorporating hesitancy data from the original apportioned weight 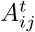 as: 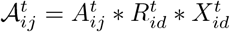

where 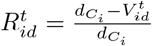 and 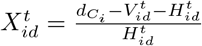

Here, 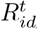 is the ratio of the unvaccinated individuals to the total population of demographic group *d* in CBG *C*_*i*_ at time *t*. 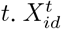 is the ratio of vaccine-accepting individuals to vaccine-hesitant individuals in the population.

Updating the apportioned weight in this approach ensures that places with high mobility are still given importance 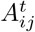, priority is reduced for CBGs where a majority of the population is vaccinated 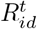, and a good balance between accepting and hesitant populations is maintained.

### 3.5 Implementation

In this section, we describe how the pipeline was implemented to support our weekly deliveries to VDH.

#### Demographic population

The demographic profiles for all the CBGs were precomputed using the Census dataset and stored as separate files in memory. Since these values are effectively static across weeks and will not change until a new census is conducted, precomputing the profiles was one way to save time during the weekly runs.

#### Model implementation

Due to the large volume of the SafeGraph Weekly Patterns data, SafeGraph issues the reports for each POI across multiple zipped files without organizing them by location or alphabetically. In order to process this large volume of data efficiently, the *upweight* and *apportion* are done on each separate file in parallel. Moreover, we filter out the CBGs and POIs that fall outside of Virginia. Association of the POIs with an L14 cell is also done during this step.

Afterwards, the output from each parallel job is concatenated into a single dataset, and the *visit counts* to each POI are aggregated by their corresponding L14 cells. Then, we generate a CSV file for each of the demographic groups containing the S2 identifier of each L14 cell, latitude and longitude of the centroid of that cell, and its visit count. The L14 cells are ranked based on their *visit count* for each county.

For each L14 cell, we also provide two additional pieces of information based on VDH’s feedback. The first is the day of the week when the L14 cell had its highest mobility; this required an additional step in the *aggregation* where we partitioned the network into the seven days of the week and aggregated them separately for each day. The second piece of information we provide is the address of the busiest POI within that L14 cell. For this, we find the POI with the maximum number of visits by summing the visits during the *aggregation* step.

Based on guidance from VDH, we are currently delivering recommendations for the target demographic groups Ethnicity Latinx (L), Race Black (B), and populations within the ages of 20 to 39 (A1), 20 to 29 (A2), and 30 to 39 (A3). We also use cumulative vaccination data from VDH to generate recommendations targeting unvaccinated (U) people. These, along with the population group “Whole population” (W) which considers foot traffic only, means we deliver recommendations for a total of seven population groups.

#### Final Output

We provide a total of nine CSV files to VDH. The first file contains all L14 locations in Virginia, along with their ranks and importance, for all target demographic groups. In a separate file, we provide the top 25 L14 locations per demographic group based on the highest aggregated mobility for each county. The rest of the seven files each contain the top 25 L14 locations per county for a specific population group.

We also provide seven HTML files, each pertaining to a specific demographic group, which displays the top 25 most visited L14 cells plotted on a map. For visualization purposes in the HTML, we use the latitude and longitude of the centroid of the L14 cell to plot its location, and annotate each site with additional information, including its rank, the demographic group it targets, and the highest-visited address of that tessellation. In Figure 1b and 1c, we display sample HTMLs generated by our model for the week of June 21 - June 27; they show recommended locations for the deployment of mobile vaccination sites in Danville City for two demographic groups. In Figure 2, we show this for five different demographic groups.

**Figure 2:**
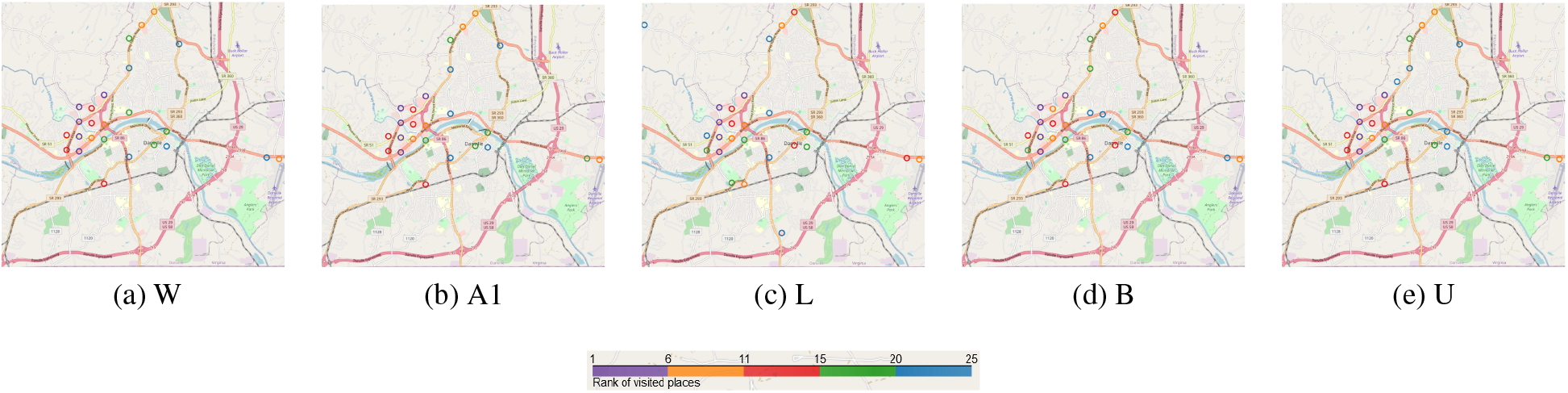
HTML maps for different demographic groups for Danville City for the week of 06-21 to 06-27. The color of the circles in the map represents the rank of the corresponding area, with lower rank meaning highly visited areas. Each circle is the centroid of a Level 14 S2 geometry.

Our model is operationalized to deliver these CSV and HTML files to VDH weekly, which VDH is currently using to target the 10 health districts with the lowest vaccination rates.

## 4 Analysis

### 4.1 Comparison of model recommendations across demographics and temporality

First, we present comparative analyses across different demographic groups at the state level. Figure 3a tabulates the number of common recommendations across the top 25 recommended sites in Virginia for pairs of demographic groups for the week of July 19 - July 25. We observe only two common recommended areas between *L* and *B*, indicating that frequently visited locations by these two demographic groups differ quite a bit. We also find that the locations frequented by *B* are quite different compared to those of the other demographic groups, as indicated by the light-colored cells across the row for *B*. Therefore, it is worthwhile to consider customized recommendations for different groups.

**Figure 3:**
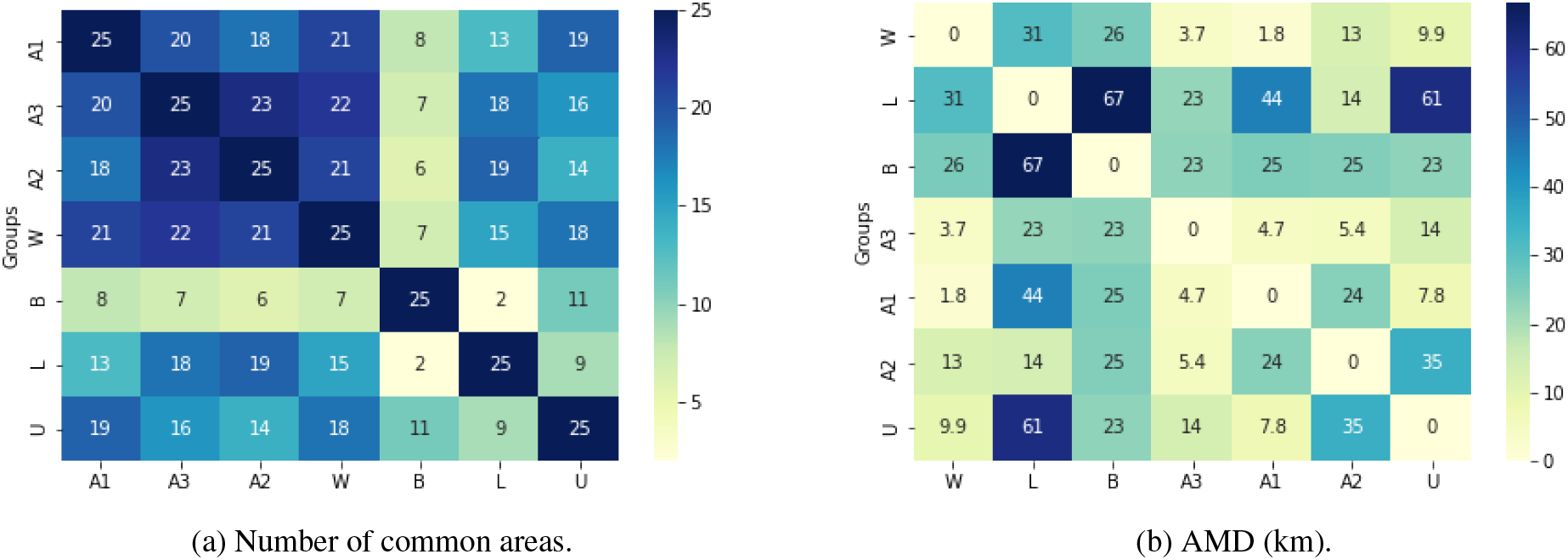
Statewide comparison of recommended places across demographics groups for the July 28 delivery.

Analyses performed across each health district also revealed similar differences between *B* and *L*, although the degree is lower than seen at the state-level. We also observe that the recommended areas are similar across different age groups for many health districts. Further discussion on this can be found in the Appendix.

Over the course of our weekly deliveries to VDH, we also noticed that recommended sites varied across different weeks. Figure 4a and 4b show variations for the top 25 recommended areas over a two-month delivery period for demographic groups *W* and *L*, respectively. Interestingly, the recommended areas are largely similar for the four weeks of June, then again across the four weeks of July. But if we compare any week in June to any week in July, the recommendations differ significantly. This suggests that mobility patterns changed after June going into the month of July. The state of emergency mandated by Virginia ended on June 30, and we wondered if this possibly contributed to this change in mobility pattern. However, when the analysis was broken down by health district, this monthly similarity pattern was less noticeable. Detailed discussion on the health district analysis is available in the Appendix.

**Figure 4:**
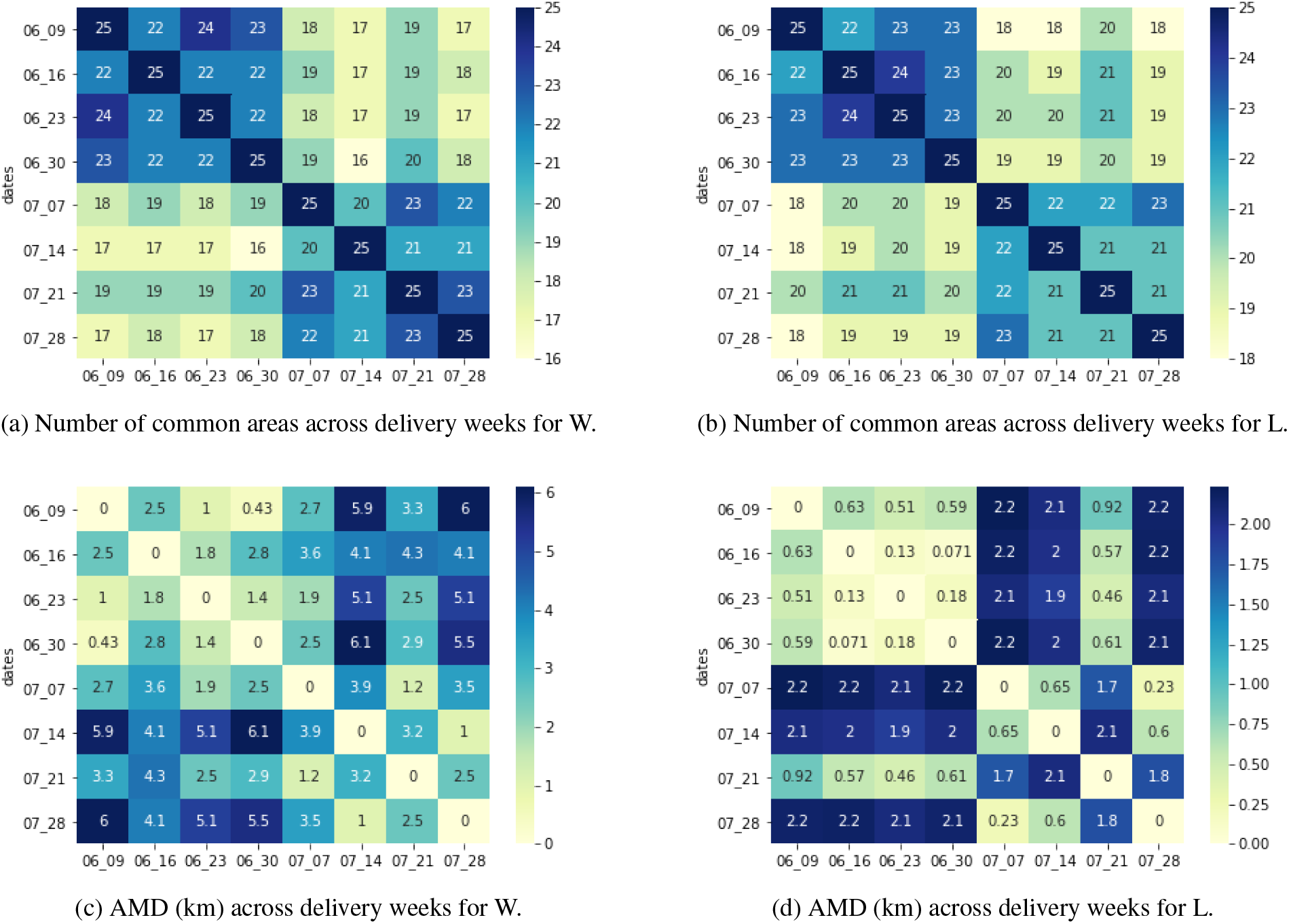
Statewide comparison of recommended places across delivery weeks.

Because the statewide phenomenon was less observed in the underlying health districts, we compared the recommended areas again from a different perspective. In studying the data, we found that the “different” L14 cells often shared a border, suggesting that aggregated foot traffic in those cells came from adjacent POIs. Therefore, instead of looking at common areas, we examined how far apart highly-ranked areas were relative to each other by calculating the average minimum distance (AMD) between two sets of areas. When comparing pairs of demographic groups for one week, the two sets are the top 25 recommended areas for those two groups. Consequently, when comparing delivery weeks for a particular group, the two sets are the top 25 recommended areas for two different weeks for a particular group.We calculated AMD by matching each area in one set with its closest area in other set and taking the average of the haversine distance of the matched areas based on their centroids.

We find that while the monthly pattern is still observable for *W* (Figure 4c), it is not the case for *L* (Figure 4d). This is contrary to the similarity observed in Figure 4a and Figure 4b. Furthermore, the AMD between areas are quite concentrated for *L* in Figure 4d while the AMD is high between areas recommended for *L* and other demographic groups (Figure 3b). This indicates that while the frequently visited locations by Latinx individuals are comparatively far away from other demographic groups, the locations themselves visited by Latinx individuals remain relatively close to each other across different weeks.

### 4.2 Synthetic Population

In this section, we compare the areas recommended for a combined demographic group with its base demographic group. Here, we use *Race Black* and *Age 20-29* as the base demographic groups and the combined demographic group represents black individuals between the ages of 20 to 29. We observe both the number of common areas and the average minimum distance when we compare the combined demographic group with the base demographic groups.

We can observe that for the combined demographic group, it has more commonality with its base demographic group of Race Black than Age 20 - 29, both in terms of location of areas (Figure 5b) and number of common areas (Figure 5a). This tells us an individual’s mobility is dominated more by their race than their age, which is somewhat intuitive given that we did not see much variation across the sampled age groups.

**Figure 5:**
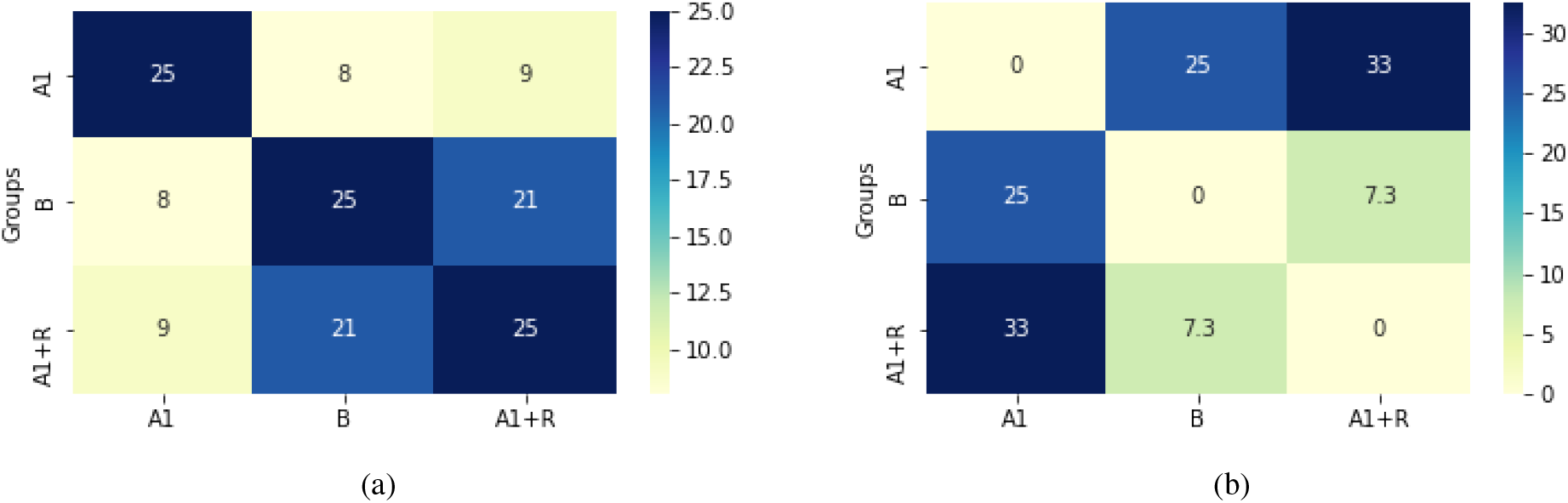
Comparison of recommended places between two base demographic groups and its combined version. The figure on the left shows the number of areas these groups have in common, and the second figure shows the average minimum distance (km) between top-ranking areas. We observe that the combined demographic group had more in common with its race aspect than with its age group in the combined demographic group.

### 4.3 Acceptance/Hesitancy

In this section, we analyze the effect of incorporating hesitancy into our model by examining the recommended mobile site placements for *B* and *L*. We refer to our base model as *M*_*B*_ and the hesitancy-incorporated model as *M*_*H*_.

First, we observe the number of common recommendations between the two implementations of the two models *M*_*B*_ and *M*_*H*_. We compare from the top 1 to the top 100 areas recommended by both models and find there are more differences across the models between the recommended areas for the group *B* than group *L* (left y-axis of Figure 6).

**Figure 6:**
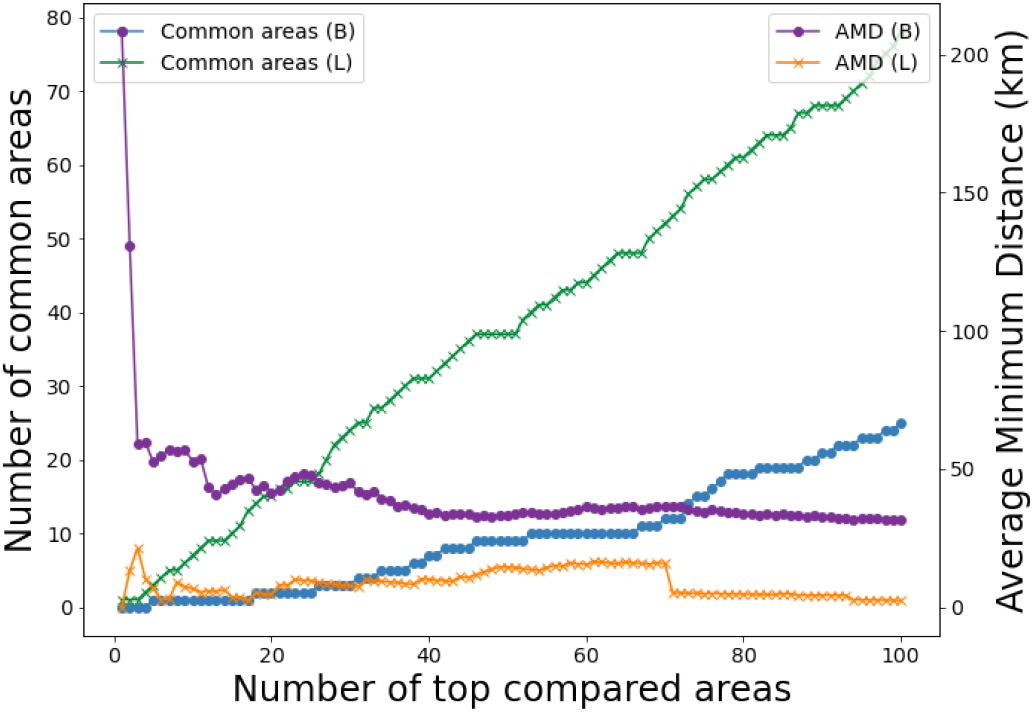
Comparison between outputs of *M*_*B*_ and *M*_*H*_.

For example, *M*_*B*_ and *M*_*H*_ have only two common areas between their recommended top 25 areas for group *B*, while there are 17 common areas for *L*. This indicates that many of the areas recommended by the base model for the Black race see high percentages of vaccine-hesitant individuals, while this phenomenon is less evident for the Latinx ethnicity group.

To explore this further, we looked at the AMD of the two sets of recommended areas considering from the top 1 to top 100 areas. We see that the two top areas for *B* in the two models are highly disparate (right y-axis of Figure 6). For example, the top recommended area by *M*_*B*_ is in Virginia Beach, which is in the Eastern region of Virginia, whereas, the top recommended area by *M*_*H*_ is in Stafford, which is in the Northern part, about 200km from Virginia Beach.

Finally, we visualize the top 100 areas recommended by both models on a map. For *B*, we see that while many areas in the southern part of Virginia are recommended by *M*_*B*_, the areas recommended by *M*_*H*_ are mostly concentrated within the Central and Northern parts of Virginia. It is also interesting that *M*_*H*_ does not recommend any area in Virginia Beach even within its top 100 recommendations, while the area was recommended highly by *M*_*B*_ (Figure 7). The model is much less sensitive to the Ethnicity Latinx group, as the map tells us that the recommended areas are mostly the same in both versions of the model (Figure 7b).

**Figure 7:**
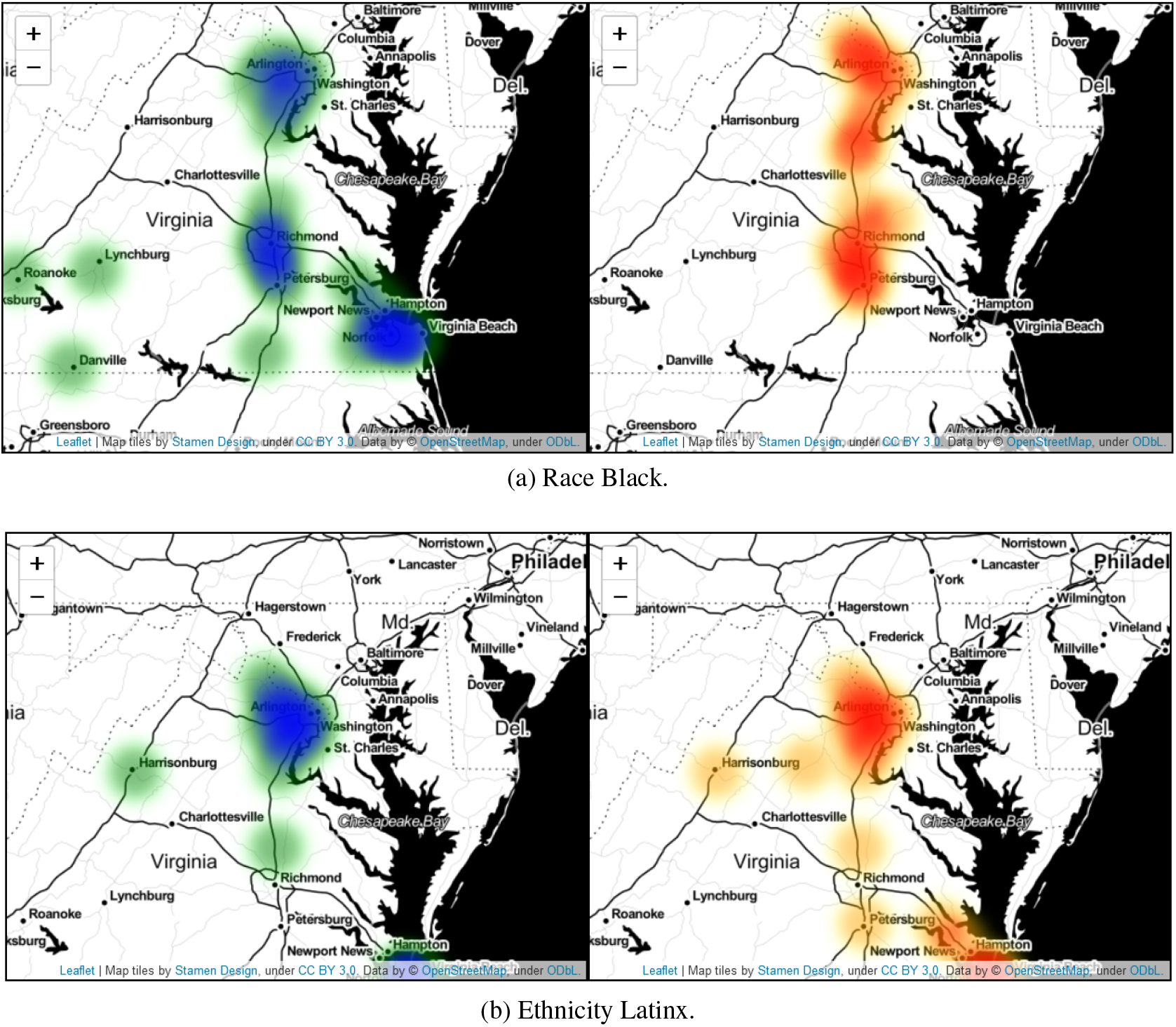
Heatmaps for areas recommended by *M*_*B*_ and *M*_*H*_. The left panel of each image shows a heatmap of the top 100 areas in the state recommended by *M*_*B*_ while the right panel is for *M*_*H*_.

### 4.4 Validation study

Evaluating the effectiveness of our strategy was a bit involved, since ground truth data indicating which of our recommended areas were used, and how accessible they were to different demographic groups, was not available. However, VDH provided a list of 147 mobile sites deployed between May 19 and June 30 along with the number of daily doses administered at those sites.

Using these sites, we conducted a retrospective analysis of the effectiveness of our strategy. We looked at the corroboration of our recommendations with these deployed sites using the recommendations for groups *B* and *L* over the month of June. Specifically, for each deployed site, we looked at our closest recommended area and found that most of the deployed sites were within 1km of at least one of our recommended areas for both groups (Figure 8a), suggesting that either our placement strategy affected these sites, or the original selection strategy and our strategy corroborated each other to some extent.

**Figure 8:**
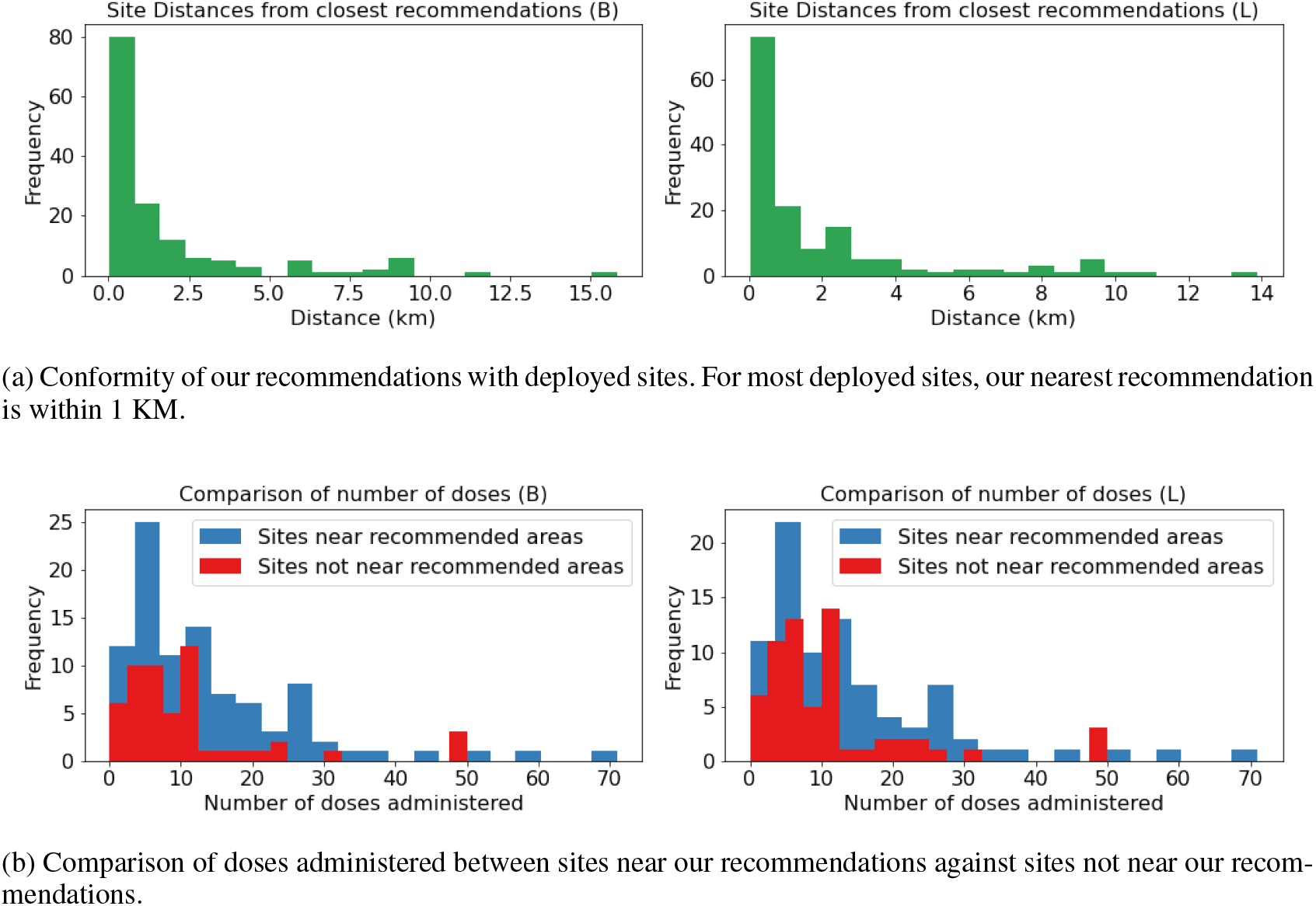
Effectiveness of our placement strategy.

It is also important to look at how effective these conforming sites were with respect to vaccine administration. For this, we compared the sites within 1km of at least one of our recommended areas with the remaining sites in terms of doses administered. We find that, in general, sites close to our recommended areas administered higher numbers of doses on average to both groups (Figure 8b). This indicates that our recommended areas are indeed more accessible, and it is probable that using our placement strategy could help policymakers to increase uptake among targeted demographic groups.

## 5 Discussion

In this work, we have devised a data-driven, equity supportive, dynamic, rule-based recommendation model for the social good that considers evolving mobility patterns to find a real-time placement strategy for making vaccines more accessible to targeted demographic groups. This model has been operational since early June 2021, and, since then, we have delivered mobile vaccination recommendations to VDH every week. Our strategy of using mobility data from SafeGraph to identify locations with high foot traffic, then refining those sites by using data from the US Census, vaccination data, and modeled vaccine acceptance data to target under-vaccinated communities, has proven effective. There are some additional takeaways from this experience that are worth noting, however.

*First*, some target demographic groups are more divergent than others. This variation demonstrates why a targeted ranking of locations is relevant, and that foot traffic alone may not be an adequate indicator for all cases. We also find that, depending on the target demographic group, incorporating hesitancy may be a necessary consideration.

*Second*, trends can appear at the state level that do not carry through to the underlying districts, which underscores the need to examine details at finer resolutions. Also, we find that while mobility may differ across different demographic groups, the mobility patterns of a particular demographic group stays comparatively stable over time.

*Third*, although our research focuses on the Commonwealth of Virginia, this approach can be generalized for any other US state. SafeGraph and the Census data have coverage across the US, and vaccine acceptance data can be modelled across the states. The targeted group that may require more work is the unvaccinated; VDH was able to provide vaccination data to census tract resolution, but county-level data may be the best we can hope for in some states.

There are some limitations to our work. It was difficult to validate our work against actual vaccination rates at the mobile vaccination sites, as that data was unavailable for most of the state; however, our site recommendations compared favorably with the sites placed by VDH where data was available. Our model also assumes that people from a given CBG who frequent these L14 locations are demographically similar to the population of that CBG; this may not hold true in all cases. In the future, we want to factor this into our model by taking into consideration that the mobility of a particular demographic group from a CBG to a POI not only depends on the demographic distribution of the CBG, but also the category of the POI.

## Data Availability

All data produced in the present study are available upon reasonable request to the authors

## 6 Acknowledgments

The authors would like to thank members of the Biocomplexity COVID-19 Response Team, Network Systems Science and Advanced Computing (NSSAC) Division, SNAP Stanford team and VDH Personnel for their thoughtful comments and suggestions. We also thank Lisa Shifflett for her help in arranging the data used for the validation study. This work was partially supported by the National Institutes of Health (NIH) Grant R01GM109718, VDH Grant PV-BII VDH COVID-19 Modeling Program VDH-21-501-0135, NSF Grant No.: OAC-1916805, NSF Expeditions in Computing Grant CCF-1918656, CCF-1917819, NSF RAPID CNS-2028004, NSF RAPID OAC-2027541, US Centers for Disease Control and Prevention 75D30119C05935, DTRA subcontract/ARA S-D00189-15-TO-01-UVA. We also gratefully acknowledge the support of DARPA under Nos. HR00112190039 (TAMI), N660011924033 (MCS); ARO under Nos. W911NF-16-1-0342 (MURI), W911NF-16-1-0171 (DURIP); NSF under Nos. OAC-1835598 (CINES), OAC-1934578 (HDR), CCF-1918940 (Expeditions), IIS-2030477 (RAPID), NIH under No. R56LM013365; Stanford Data Science Initiative, Wu Tsai Neurosciences Institute, Chan Zuckerberg Biohub, Amazon, JPMorgan Chase, Docomo, Hitachi, Intel, KDDI, Toshiba, NEC, and UnitedHealth Group.

J.L. is a Chan Zuckerberg Biohub investigator.

Any opinions, findings, and conclusions or recommendations expressed in this material are those of the author(s) and do not necessarily reflect the views of the funding agencies.

## 7 Ethics Oversight

This study and the data sources used for this study were evaluated by the Institutional Review Board for Social and Behavioral Sciences (IRB-SBS) at the University of Virginia; the IRB-SBS Chair is Tonya Moon, and the IRB-SBS Director is Bronwyn Blackwood. The Protocol Number is 4206, and the Protocol Title is “Biocomplexity Institute COVID-19 Response: Surveillance, Modeling, and Operational Support”. This protocol was most recently approved by the IRB-SBS on 2021-10-15.

## A Southside Pilot Study

In this section, we describe a baseline study which helped inform the development of our final model.

In this study, we recommended placement of mobile vaccine clinics serving the Black and Latinx populations in the Southside Health District of Virginia using traditional location-allocation methodologies as provided by commercial Geographic Information System Mapping (GIS) software. The goal of these analyses was to select a subset of provided candidate sites that would collectively minimize travel impedance from demand points Tomintz et al. (2015). Given the flexibility of the mobile vaccine clinics, our candidate facilities included any large commercial or government building, or parking lot in the district. Specifically, candidate sites included all public schools as provided by the Virginia Department of Education, box stores, grocers, movie theaters, sporting arenas, and public buildings, including fire stations, Emergency Medical Services (EMS) stations, libraries, and government centers found in the OpenStreetMap points-of-interest dataset Haklay et al. (2008). Candidate sites within 500 meters of each other were clustered using DBScan and reduced to their collective centroids, resulting in 86 unique entries. Households with at least one Black or Latinx resident served as demand points, and each household was weighted by the number of such residents as based on data derived from the Biocomplexity Institute’s synthetic population, which includes individual households with attached demographic data at the street address level Mortveit et al. (2020); this is described in more detail in Section 3.1. Households were clustered using a 200-meter DBScan, and their weights were aggregated, resulting in 12,441 demand points representing 26,906 individuals.

Location allocations were done using Esri ArcGIS Pro 2.8 and the ArcGIS REST API to calculate and minimize drive times. As the health district is quite rural and there is minimal public transportation, we did not consider alternative modes of travel. We calculated two solutions of 20 vaccine clinics each, one minimizing population-weighted travel time, the other maximizing attendance on the assumption that facility utilization would be inversely proportional to travel time squared. The former, often called the “P-median” problem, is common in healthcare facility placement Polo et al. (2015); Rahman et al. (2000), while the latter is based on the gravity model of spatial access Wan, Zou, and Sternberg (2012); Lowe et al. (1996) and has shown application in predicting healthcare-related distance decay Stulz et al. (2018). The results were submitted for review by domain experts within the district and compared to existing VDH-selected sites. The results were also used as a baseline for future mobility-based clinic placements.

Through this study, it became evident that areas with high foot traffic could be potential candidates for vaccine distribution sites. However, instead of choosing only a few categories of POIs, it may have been worthwhile to explore a more diverse set of POI types. Furthermore, variation was seen in the most visited places across different demographic groups.

## B Model with foot traffic only

In this model, we recommend areas based on overall foot traffic to SafeGraph’s POIs without considering demographic information. This process was as follows:

1. We found all the S2 Geometry L8 cells inside the state of Virginia. These L8 cells were approximately county-sized and are defined as *regions* for this model.
2. Subsequently, we divided each L8 cell into its corresponding S2 Geometry L14 cells. These L14 cells are defined as *areas*. These areas usually contained no more than a dozen POIs.
3. We identified the areas with the most foot traffic for each region by aggregating the sum of the visits to its underlying POIs.

For each L8 cell and day of the week, we reported the top 10 highest-mobility L14 cells. Let *P*_*l*_ represent the set of POIs located in a given L14 cell *l*. Then, we computed *l*’s foot traffic, 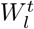, on a given day *t* by summing over the total foot traffic to any POI in *P*_*l*_:

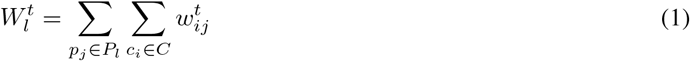

To identify the highest-mobility areas for each day of the week, we summed over data for that day from the most recent three weeks of SafeGraph’s Weekly Patterns data (which were April 26 to May 16, 2021, at the time when we conducted this part of the study). We chose to only use data from the most recent weeks, since mobility patterns were rapidly changing due to lifting of COVID-19 restrictions and widespread vaccination.

## C Vaccine Hesitancy Simulation Equations

Tracking vaccine acceptance and hesitancy is not a straightforward endeavor. While surveys may be employed to measure hesitancy, it is difficult to conduct a timely survey that can accurately assess acceptance among different demographic groups within a small geographic area. We instead employ a simulation which models vaccine hesitancy relative to vaccine uptake. For a given population of *n* individuals, all persons are in one of three states: hesitant *H*, accepting *A*, or vaccinated *V*. The model dynamics are explained thoroughly below:

Let *V*_*t*_ be the number of people within the population who are vaccinated at time *t*, let *A*_*t*_ be the number of people in the population who are accepting of the vaccine at time *t*, and let *H*_*t*_ be the number of people in the population who are hesitant. For all *t, V*_*t*_ + *A*_*t*_ + *H*_*t*_ = *n*. We make the further assumption that people move between these states in fixed proportions as follows.

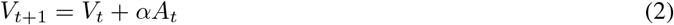

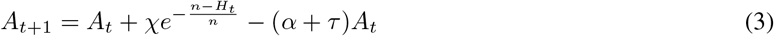

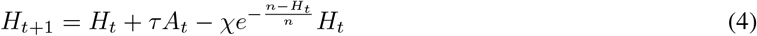

In other words, people who are accepting of the vaccine become vaccinated at some rate *α*. People who are accepting either become vaccinated at rate *α*, or become hesitant at rate *τ*. People who are hesitant become accepting at a rate proportional to 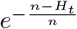. This models “polarization”; where the fewer people who are hesitant, the less likely it is that the hesitant become accepting.

This simulation is parameterized by *α, τ, χ*, and the initial conditions *V*_0_, *A*_0_, *H*_0_. We can empirically observe *V*_0_ for each county via vaccine administration data. We assume that *α* (vaccine accessibility) and *A*_0_ (initial acceptance) vary from county to county, whereas the *χ* and *τ* are constant across all of Virginia. We find the best simulation by enumerating a grid of values, and selecting the simulation that best fits the observed vaccine uptake over all *t* and *p*. The parameter space we used is *chi, τ* ∈ [0.05, 0.8], *α* ∈ [0.1, 0.4], and *A*_0_ ∈ [0.3, 0.8].

After finding the best parameters, we then calibrate a simulation for each demographic group. The *R*^2^ value of our simulation’s vaccine uptake against empirically observed vaccine uptake was 0.94 for Black populations and 0.99 for Latinx populations across all Virginia counties for the time period considered.

## D Analysis

### D.1 Comparison of model recommendations by demographics and temporality across health districts

Figure 9 shows the pairwise demographic analyses performed for each health district. The difference in the top 25 recommended sites between the Black race (B) and other demographic groups can again be observed for some health districts, although the difference is lower compared to the state-level analysis. Additionally, we find that across some districts, the Latinx ethnicity group (L) also displays this kind of disparity. Another observation is that the top 25 recommended places are largely similar for different age groups across most of the VDH health districts. This is apparent from the dark 3*X*3 cluster formed in the top left region of the tables for many health districts (e.g. Cumberland Plateau, Henrico, and Fairfax).

**Figure 9:**
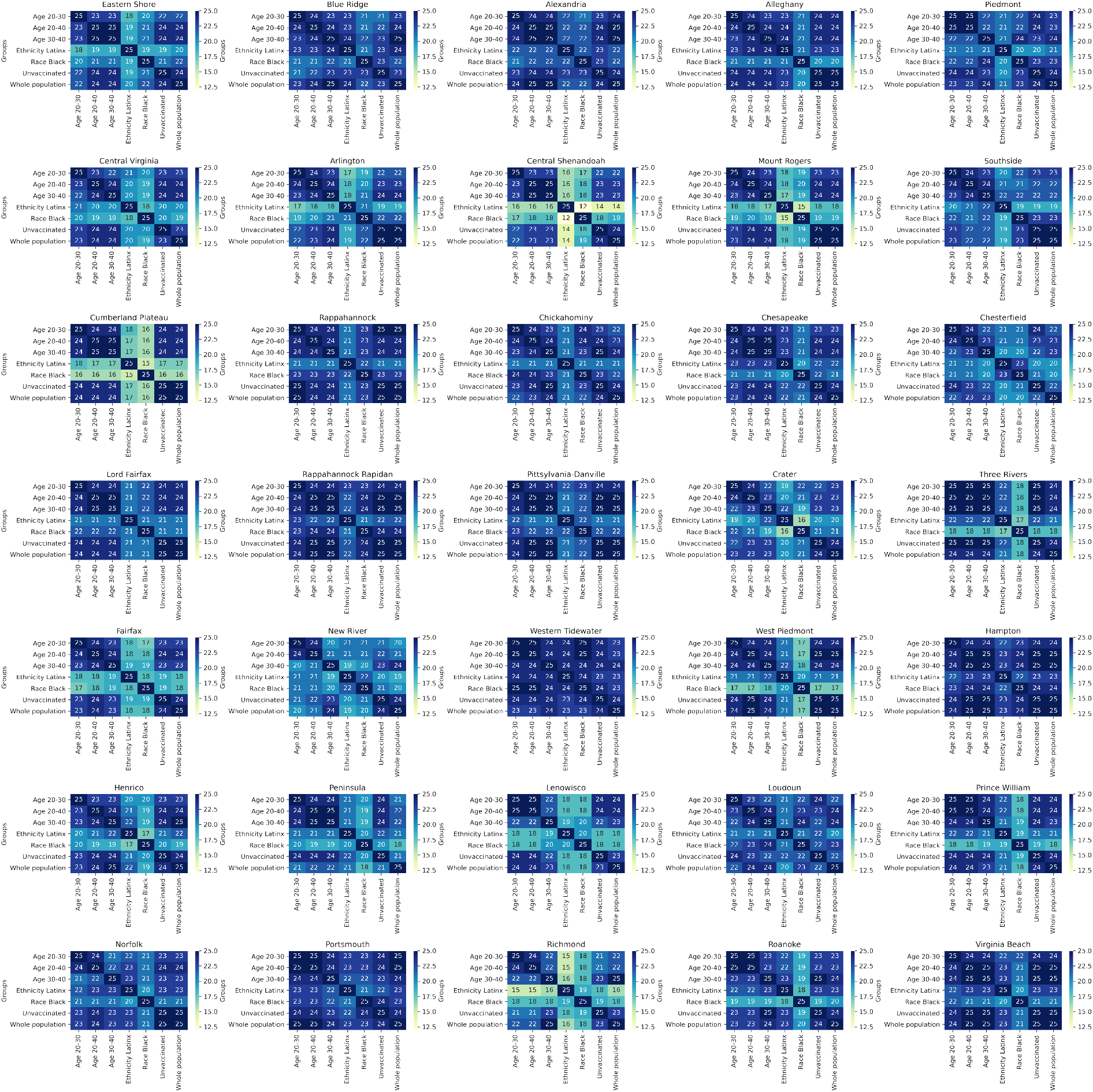
Common recommendations in the top 25 recommended sites per health district across different demographic groups.

Figure 12 shows the same analysis in terms of average minimum distance. We find that across a lot of health districts, the top recommended areas are different for *B* than other demographic groups (e.g. Roanoke, West Piedmont). However, one thing to note is that the AMD values are lower (mostly within 1 KM) than what was observed at the state-level.

Figure 10 shows the analysis across different delivery weeks. A few health districts show the same monthly similarity pattern for June and July as we saw at the state level (e.g. Henrico), while some health districts show the pattern for only one month (e.g. Chesterfield, Mount Rogers). Other health districts do not show the same similarity pattern across the monthly divide. In figure 11, we make the same analysis for *L*. The monthly pattern diminishes even more. However, we do see patches of tri-weekly patterns (e.g. Three Rivers, Prince Williams) in some health districts indicating there is some correlation between the results for the two demographic groups.

**Figure 10:**
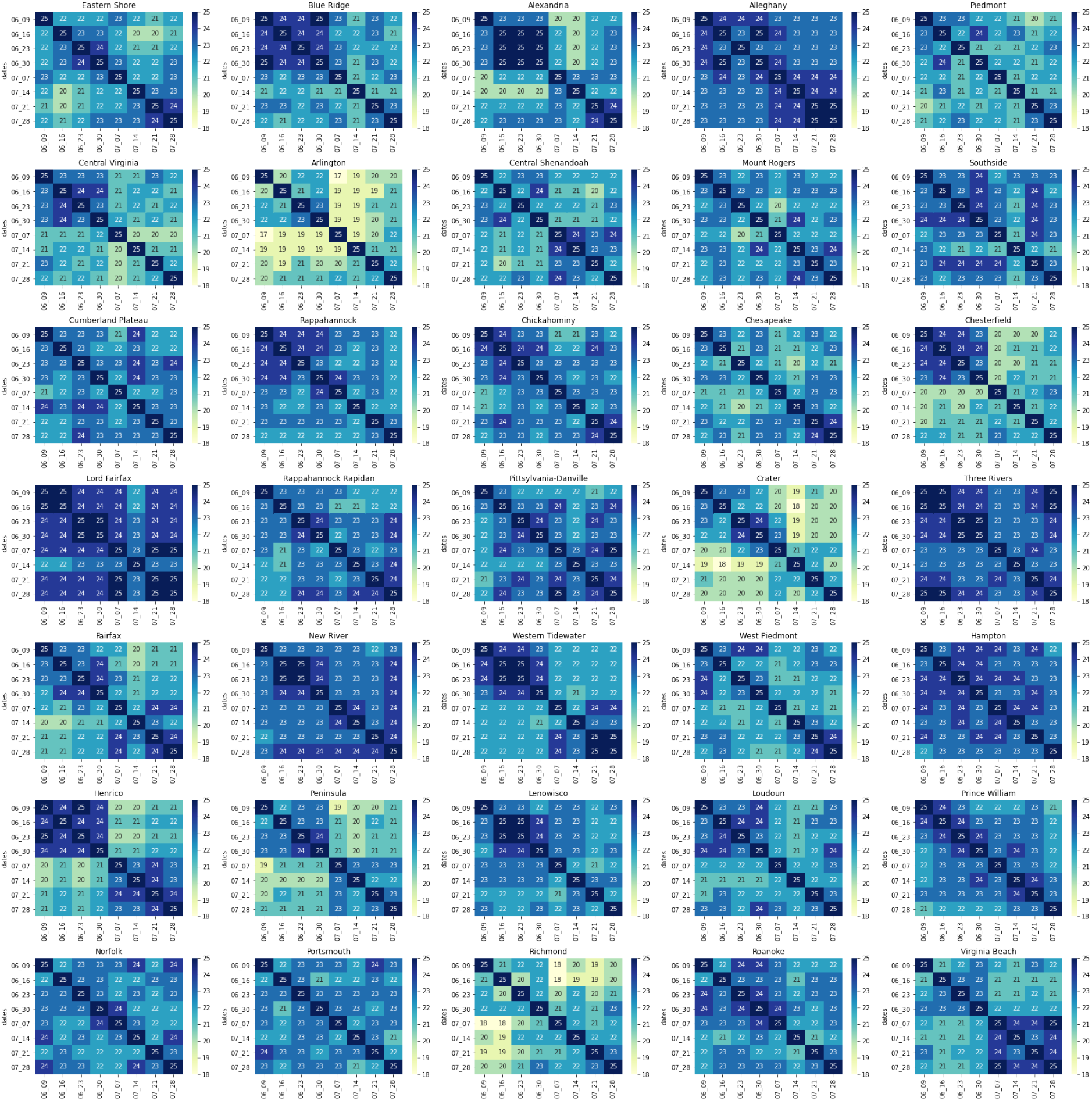
Common recommendations in the top 25 recommended places per health district across different delivery dates for *W*.

**Figure 11:**
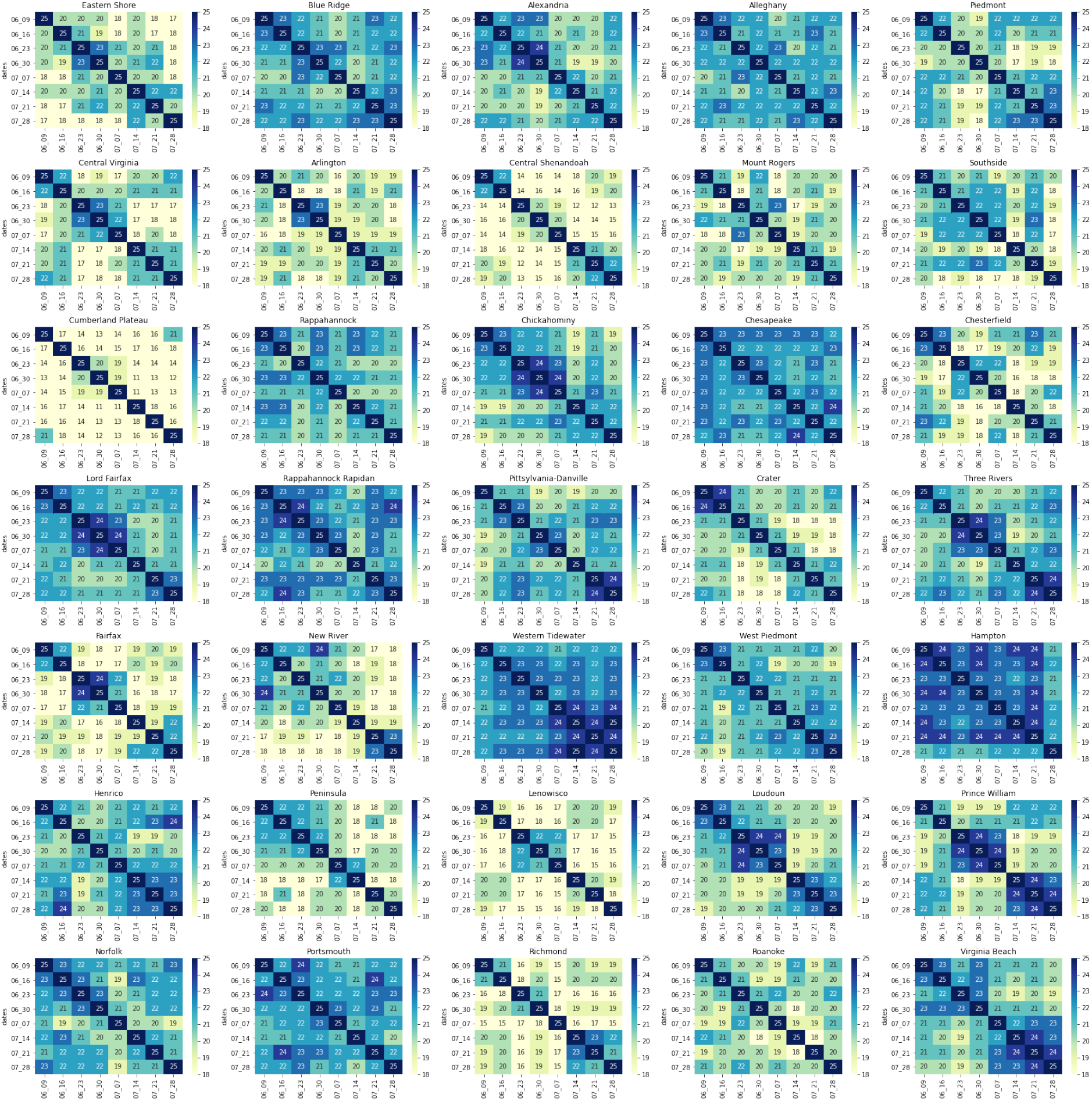
Common recommendations in the top 25 recommended places per health district across different delivery dates for *L*.

**Figure 12:**
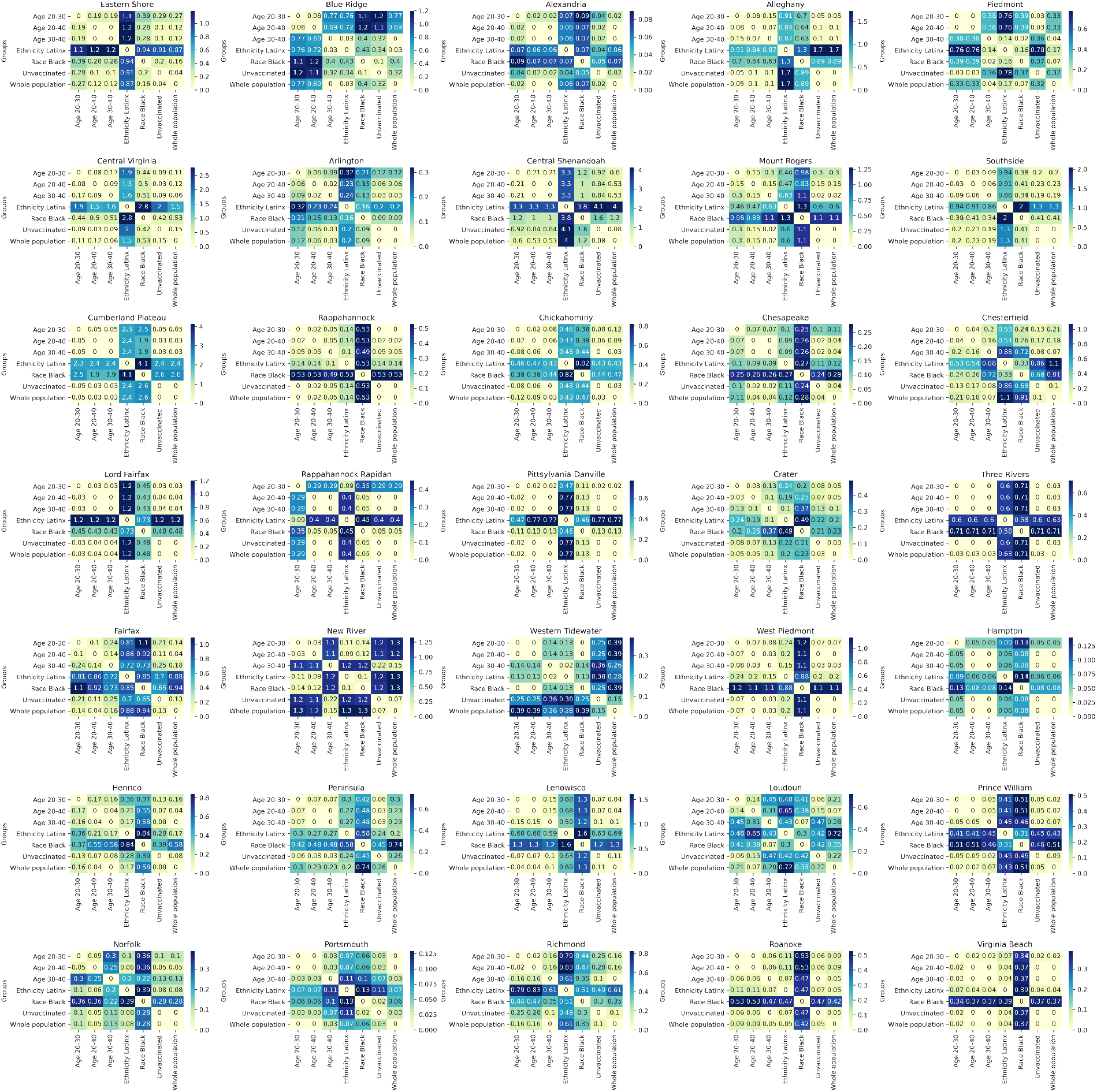
AMD between the top 25 recommended sites across different demographic groups per health district.

Figures 13 and 14 show the analysis across different delivery weeks in terms of AMD. We see that the monthly pattern is totally absent for Whole Population (W). In fact, the recommended areas are close to each other in most health districts apart from a few exceptions (e.g. Blue Ridge, Piedmont, and Crater). For *L*, no clear pattern is observed across most of the health districts, indicating the mobility pattern is quite diverse for this group and depends upon the locality.

**Figure 13:**
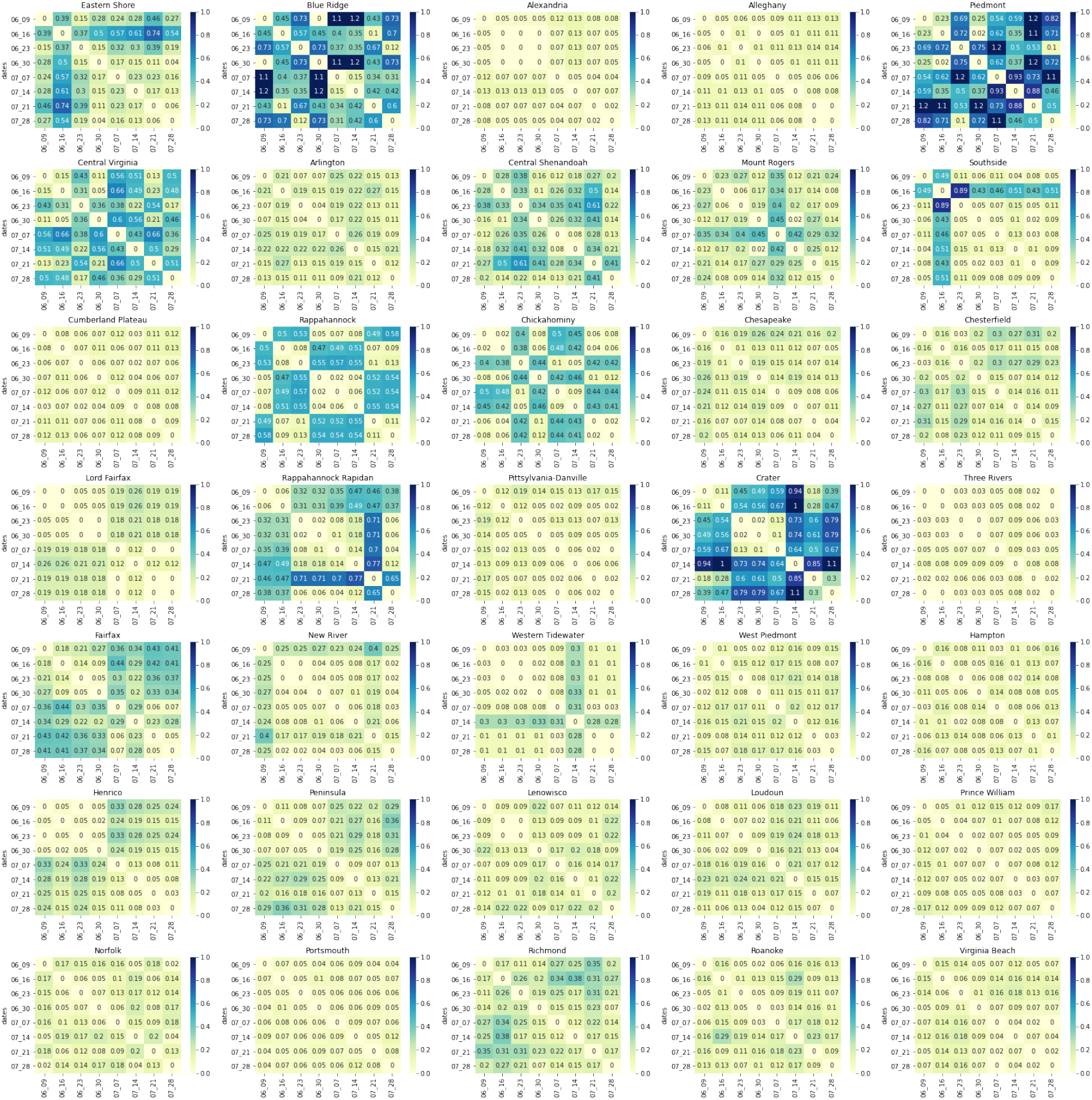
AMD between the top 25 recommended sites across delivery weeks per health district for *W*.

**Figure 14:**
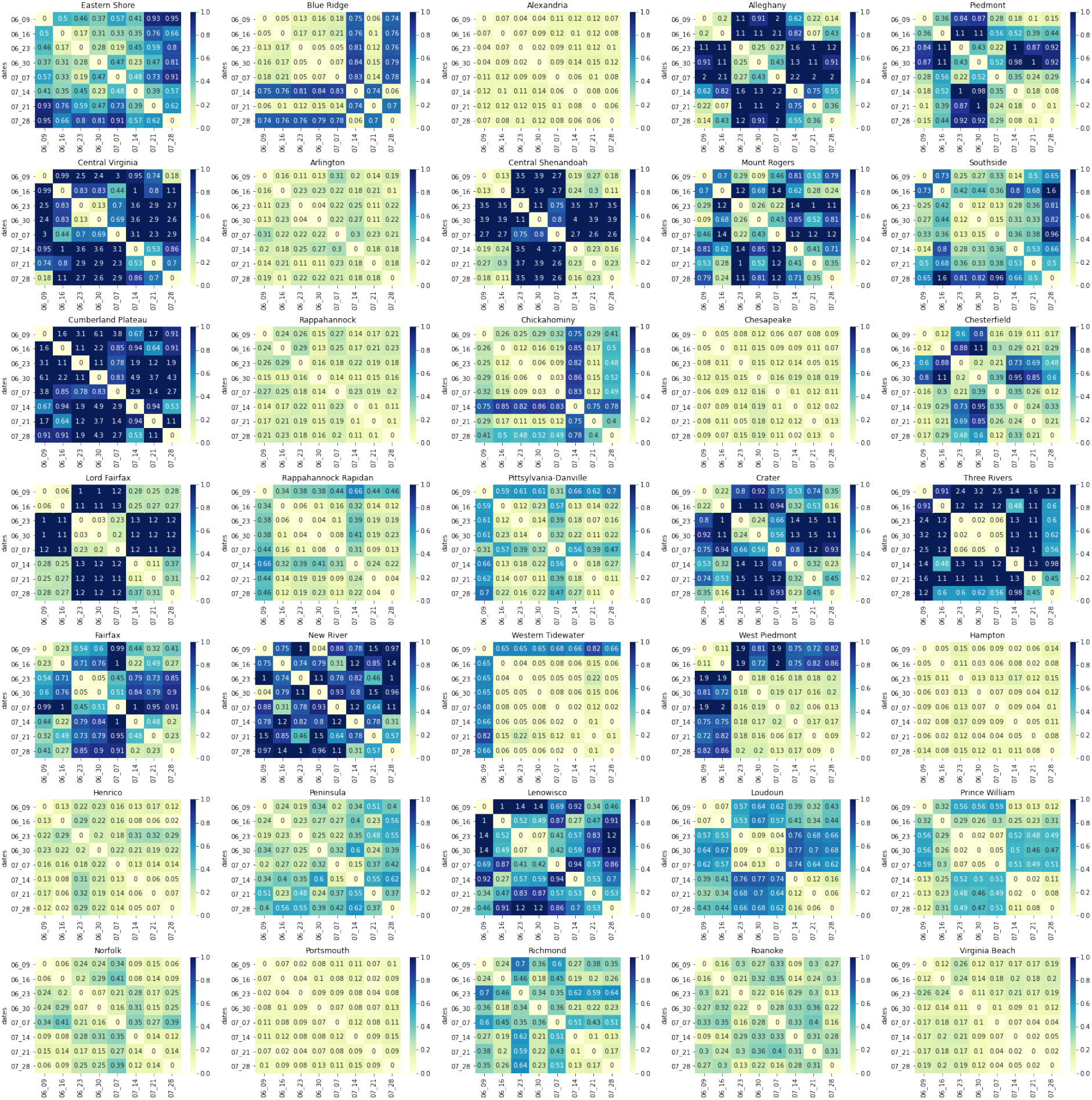
AMD between the top 25 recommended sites across delivery weeks per health district for *L*.

## Footnotes

1 https://docs.safegraph.com/v4.0/docs/weekly-patterns

2 https://s2geometry.io/

## Notes

### Competing Interest Statement

The authors have declared no competing interest.

### Author Declarations

Institutional Review Board for Social and Behavioral Sciences of the University of Virginia gave ethical approval for this work on 2021-10-15 under Protocol #4206, titled "Biocomplexity Institute COVID-19 Response: Surveillance, Modeling, and Operational Support"

